# Predictors of Healthcare Costs among Commercially-Insured Persons with Tuberculosis in the United States, 2013 to 2018

**DOI:** 10.64898/2026.08.25.26361351

**Authors:** Vinith Ilavarasan, Matthew Murrill, Robert J. Wong, Amit S. Chitnis, Devan Jaganath

**Affiliations:** UC Berkeley School of Public Health, Berkeley, California; Division of Hospital Medicine, Department of Medicine, University of California, San Francisco, San Francisco, California; Center for Tuberculosis, University of California, San Francisco, San Francisco, California; Division of Gastroenterology and Hepatology, Stanford University School of Medicine, Palo Alto, California; Tuberculosis Section, Division of Communicable Disease Control and Prevention, Alameda County Public Health Department, San Leandro, California; Division of Pediatric Infectious Diseases, Department of Pediatrics, University of California, San Francisco, San Francisco, California

**Author notes:** **Corresponding Author:** Devan Jaganath, MD, MPH, Division of Pediatric Infectious Diseases, University of California, San Francisco, 550 16th Street, 4th Floor, San Francisco, CA 94158,).

**Keywords:** tuberculosis, health expenditures, hospitalization, insurance, health, public health

## Abstract

**Context:** Most individuals in the United States have commercial health insurance, yet costs for tuberculosis (TB) care have focused on the public sector.

**Objective:** To quantify 12-month all-cause healthcare costs and identify predictors of expenditure among commercially insured persons with TB disease in the United States.

**Design/Setting:** Retrospective cohort study using Merative™ MarketScan® Commercial Claims Database (2013–2018).

**Participants:** Adults ≥18 years old with TB disease

**Main Outcome Measure:** Total 12-month all-cause healthcare costs (outpatient, inpatient, pharmacy) were calculated from the date of diagnosis. Adjusted cost ratios (aCR) were estimated using a Gamma generalized linear model.

**Results:** We included 303 individuals diagnosed with TB disease, median age 46 years, 158 (52%) male, 16 (5%) with HIV, 12 (4%) with hepatitis B (HBV), and 13 (4%) with a drug use disorder. Mean total 12-month costs were $32,404 (median $8,075; SD $78,829). Median 12-month costs were substantially higher among persons with any comorbidity (HIV, HBV, hepatitis C (HCV), alcohol use disorder, drug use disorder, or Charlson score >0) compared to those without ($11,930 [IQR $4,194–$36,073] vs $3,385 [IQR $1,506–$8,609]; p<0.001). HIV coinfection and drug use disorder were the strongest independent predictors. HIV coinfection was associated with 4.7-fold higher costs (aCR 4.70, p<.001), driven predominantly by pharmacy expenditure (aCR 16.4). Drug use disorder was associated with 3.2-fold higher costs (aCR 2.62, p=.03). Comorbidity burden was a continuous independent predictor (aCR 1.36 per Charlson point, p<.001).

**Conclusions:** Healthcare costs are high among persons with TB who have commercial insurance, and are further increased with comorbidities including HIV coinfection and drug use disorder. Improved screening, care coordination and management of TB and high-risk comorbidities could yield significant cost savings.

## INTRODUCTION

For the last four decades, the public sector has played a significant role in supporting tuberculosis (TB) diagnostic and treatment efforts in the United States (U.S.).^1^ There has, however, been a substantial shift towards private sector payment. Health-department–managed TB cases declined steadily from 1993–2006, while the private-sector share of paid cases rose from 25% to 32% between 2007 and 2011.^2^In addition, the signing of the Patient Protection and Affordable Care Act in 2010, signaled a potential furthering of this public-to-private shift of domestic TB payer burden, given that millions of previously uninsured Americans were brought into the private insurance system.^3^ As a result, the proportion of persons diagnosed with TB in the United States who receive care through commercial health insurance is presumably also increasing. This makes understanding both the healthcare costs and resource requirements of this population critical for resource allocation.

Researchers have investigated the cost of hospitalization^4^ among this increasingly privately insured TB population. It was found that, between 2010 and 2014, average hospitalization expenditure per privately insured person was $33,085^4^ as compared to the all-payer average of $16,695.^5^ However, most TB care occurs primarily in the outpatient setting, and hospitalization represents only a portion of total healthcare costs. A critical gap exists in understanding the all-cause healthcare costs of commercially insured persons with TB, including not only inpatient costs, but also outpatient and pharmacy expenditures. Furthermore, comorbidities including HIV^6^, substance use disorder^7^, and hepatitis B^8^ play a significant role in TB disease severity, treatment complexity, and clinical outcomes. However, limited research has been conducted on the impact of these comorbidities on overall healthcare costs, particularly among commercially insured populations.

To address this evidence gap, we conducted a retrospective cohort study of commercially insured persons diagnosed with TB using the MarketScan Commercial Claims Database (2013-2018). The objectives of the study were: 1) to quantify the 12-month all-cause healthcare costs (including outpatient, inpatient, and pharmacy) for patients with TB disease in the commercial insurance setting, 2) to identify patient-level predictors of healthcare expenditure, and 3) to examine the role of comorbidities in driving TB-related healthcare costs.

## METHODS

### Data Source and Study Population

For this study, private insurance claims data for reimbursement from the Merative™ MarketScan® Commercial Claims Database (2013–2018).^9^ These claims are exclusively from the under-65 population and contain de-identified claims from over 300 large, self-insured U.S. employers, health plans, and managed care organizations.^9^ The databases contain data from outpatient drug claims, outpatient services, inpatient services, and inpatient admissions from 2013 to 2018. For this study, only adults with at least one pharmacy claim during the study period were included.

Analysis was restricted to persons with TB disease to remain consistent with previous TB cost studies.^4,10^ While mycobacterial culture^11^ is the established gold standard for the laboratory diagnosis of TB disease, claims data containing culture orders/results were not available.

Because prior studies have noted limited validity of TB case definitions based on International Classification of Diseases (ICD) diagnostic codes alone,^12^ we used a validated case definition for TB that incorporates both ICD codes and prescription claims.^13^ Persons met our primary case definition if they had both: 1) a pharmacy claim for pyrazinamide on any date in the study period or prescriptions of isoniazid and rifampin filled on the same day and 2) at least two TB-coded encounters on different service date.

The index TB diagnosis date was defined as the later of a) the first date the medication criterion was met or b) the date of the qualifying second tuberculosis-coded encounter. This date represents the time at which both components of our identification criteria were met.

All persons with a TB diagnosis were included in the analyses if they had continuous plan enrollment for at least six months before and 12 months following the index date. Index dates were also restricted to the period of July 2013 to December 2017 to ensure full enrollment windows were observable within the 2013-2018 data span.

Each patient contributed only one index date, defined by their first qualifying TB episode. Patients with evidence of prior or recurrent TB disease (defined as earliest TB-related claims occurring more than 365 days before the index date (n=16, 5.3%)) were retained in the primary analysis but excluded in a pre-specified sensitivity analysis to assess whether recurrent cases influenced findings.

### Exposures, comorbidities, and predictor definitions

The Charlson Comorbidity Index (CCI) is a validated tool used to predict 10-year survival of weighting 17 different comorbidities.^14^ We identified comorbidities by filtering inpatient and outpatient diagnosis codes in the six-month pre-index lookback window for each patient (**Appendix A**). We used a previously published, standardized algorithmic approach for claims-based Charlson scoring in United States health services research.^15^

HIV was not included in the Charlson composite score so as not to double-count. HIV, HCV, alcohol use, and substance use were defined as >1 comorbidity-related ICD code in the six-month lookback window. TB disease site was also classified hierarchically from TB ICD codes: pulmonary-only; extrapulmonary-only; or both pulmonary and extrapulmonary (codes from both groups present). Drug-induced liver injury (DILI) was identified using ICD-9 codes for toxic hepatitis and ICD-10 codes K71.0 through K71.9. We classified DILI events as pre-existing (within the 6-month period prior to diagnosis date), or incident (occurring during the 12-month follow-up period without any pre-existing DILI claim). ICD-9 and ICD-10 codes for each comorbidity were identified based on the code lists published by the Centers for Medicare Services.^16^

### Cost measurement

We measured all-cause healthcare costs over the 12 months following the index date, capturing outpatient services, inpatient admissions, and outpatient pharmacy fills. Outpatient and pharmacy costs were summed from the gross payment field on each claim, representing the total amount paid to the provider before patient cost-sharing. Inpatient costs were summed from the total case payment field on admission records, which consolidates facility and physician payments for each hospitalization. Emergency department visits were identified using the standard place-of-service code for hospital emergency rooms. TB-specific medication costs were computed as the subset of pharmacy costs attributable to the four first-line antitubercular drugs: isoniazid, rifampin, pyrazinamide, and ethambutol.

All costs were inflated to 2018 U.S. dollars using the Medical Care component of the Consumer Price Index (Bureau of Labor Statistics).^17^

All-case costs were selected over TB-attributable costs only because the objective of this study was to identify predictors of healthcare expenditure in commercially insured persons with TB, including expenditures attributable to coinfections, comorbidities, and treatment-related complications that may not be coded as TB-related, but represent the real costs of TB care.

### Statistical Analysis

Healthcare cost distributions were right-skewed and strictly positive, consistent with prior TB cost analyses. Total twelve-month costs were modeled using a generalized linear model with a Gamma distribution and log link function, as recommended by Basu and Manning for healthcare cost outcomes^18^.

The primary multivariable model included the following predictors: HIV coinfection, Drug use disorder, Alcohol use disorder, Incident DILI, modified Charlson Comorbidity (continuous, per point), TB disease site, HBV coinfection, age category, sex, U.S. Census region where the person received care, and calendar year of index TB diagnosis date (2013 through 2017).

To assess whether the association between Charlson score and costs was approximately linear on the log scale, we examined model residuals by Charlson score category and refitted the primary model treating Charlson score as a categorical variable (0, 1, 2, ≥3). Residuals showed no systematic pattern across Charlson categories, and the categorical model produced consistent findings for all other predictors, supporting the use of the continuous specification.

To examine where each predictor’s cost effect was concentrated across the care continuum, parallel models were fit for outpatient costs (Gamma GLM, log link), inpatient costs among admitted patients only (Gamma GLM, log link), outpatient pharmacy costs (Gamma GLM, log link), and the odds of any inpatient admission (logistic regression). Because inpatient expenditure was zero for patients without an admission, we modeled the probability of hospitalization separately using logistic regression and conditional inpatient costs among hospitalized patients using a Gamma GLM..

### Sensitivity Analyses

Three pre-specified sensitivity analyses were conducted. The primary model was refit using an expanded 15-month cost window (90 days pre-index through 365 days post-index) to capture pre-confirmation diagnostic encounters. We also refit the model excluding the four persons with costs exceeding the 99th percentile ($412,922) and separately excluding the 16 patients with the earliest TB-related claims more than 365 days before their index date, who likely represented prior or recurrent diagnoses.

### Software and reproducibility

All analyses were conducted in R version 4.1.1 base R glm() for regression modeling. Statistical significance was defined as a two-sided p-value <0.05.

### Ethics

This study used de-identified secondary data from a commercial claims database and was determined to be exempt from review by the UCSF Institutional Review Board.

## Results

### Participant Flow and Characteristics

Of the 9,586 commercially insured adults with at least one pharmacy claim from 2013-2018, 826 met our medication criterion (pyrazinamide on any date, or simultaneous isoniazid and rifampin fills); 820 of these persons also had at least one TB-coded encounter (**Figure 1**). Of these, 772 met the case definition. After applying the requirements for 6 months of pre-index and 12 months of post-index continuous enrollment, 303 persons were included in the primary analytic cohort.

**Figure 1.**
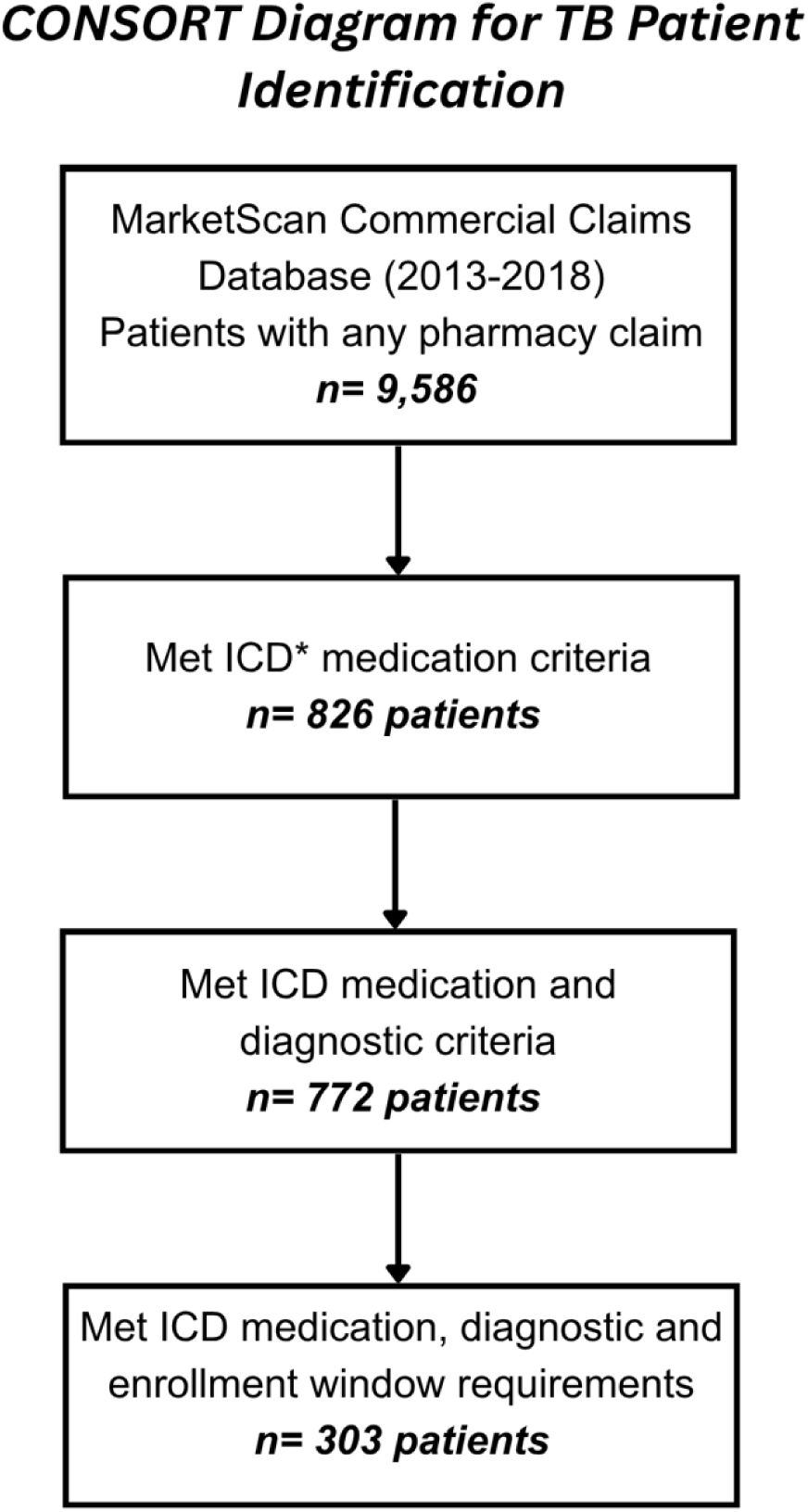
Consort Diagram for the Identification of persons with TB disease diagnoses in MarketScan Commercial Claims Database, 2013-2018. *ICD: International Classification of Diseases Persons met our primary case definition if they had both: 1) a pharmacy claim for pyrazinamide on any date in the study period or prescriptions of isoniazid and rifampin filled on the same day and 2) at least two TB-coded encounters on different service date.

The median age of the cohort was 46 years and 158 were male (52%) (**Table 1**). The cohort was geographically distributed across the four U.S. census regions. The largest representation came from the South (31%) and West region (30%). Pulmonary-only TB accounted for 60% of cases (n=182), extrapulmonary-only TB for 14% (n=42), and combined pulmonary and extrapulmonary disease for 26% (n=79).

**Table 1.**
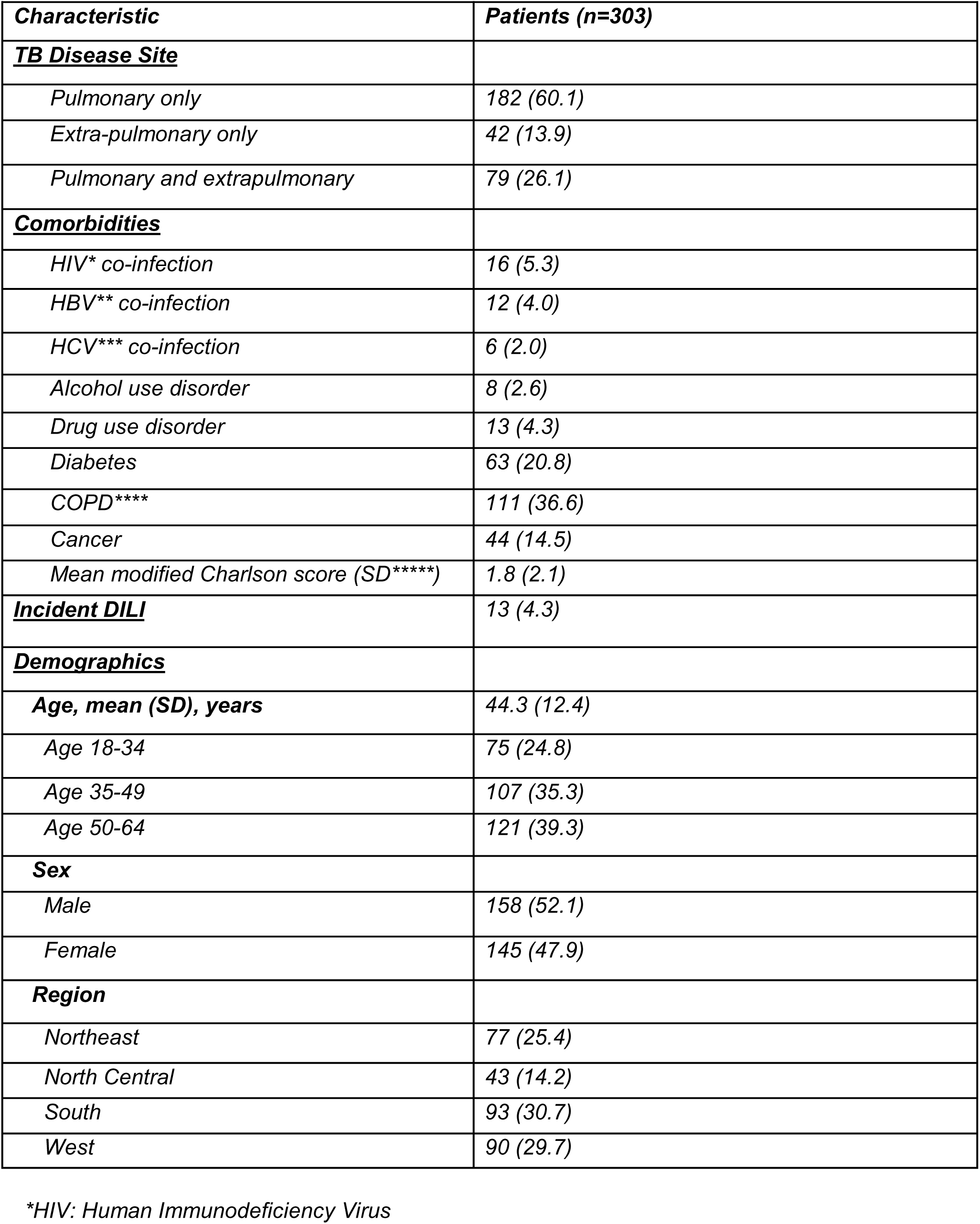
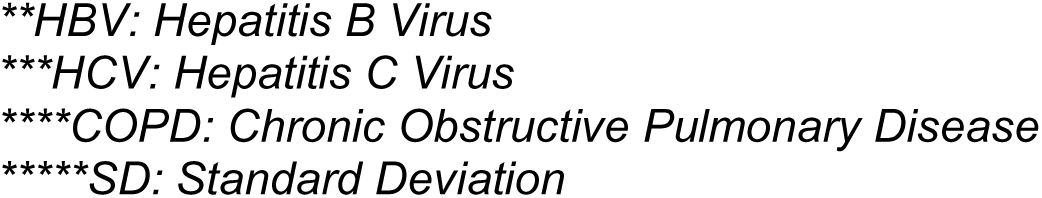
Baseline Characteristics of Persons with TB Disease.

| **Characteristic** | **Patients (n=303)** |
| --- | --- |
| **<u>TB Disease Site</u>** |  |
| *Pulmonary only* | 182 (60.1) |
| *Extra-pulmonary only* | 42 (13.9) |
| *Pulmonary and extrapulmonary* | 79 (26.1) |
| **<u>Comorbidities</u>** |  |
| *HIV\* co-infection* | 16 (5.3) |
| *HBV\*\* co-infection* | 12 (4.0) |
| *HCV\*\*\* co-infection* | 6 (2.0) |
| *Alcohol use disorder* | 8 (2.6) |
| *Drug use disorder* | 13 (4.3) |
| *Diabetes* | 63 (20.8) |
| *COPD\*\*\*\** | 111 (36.6) |
| *Cancer* | 44 (14.5) |
| *Mean modified Charlson score (SD\*\*\*\*\*)* | 1.8 (2.1) |
| **<u>Incident DILI</u>** | 13 (4.3) |
| **<u>Demographics</u>** |  |
| ***Age, mean (SD), years*** | 44.3 (12.4) |
| *Age 18-34* | 75 (24.8) |
| *Age 35-49* | 107 (35.3) |
| *Age 50-64* | 121 (39.3) |
| ***Sex*** |  |
| *Male* | 158 (52.1) |
| *Female* | 145 (47.9) |
| ***Region*** |  |
| *Northeast* | 77 (25.4) |
| *North Central* | 43 (14.2) |
| *South* | 93 (30.7) |
| *West* | 90 (29.7) |

\*HIV: Human Immunodeficiency Virus
**\*\*HBV: Hepatitis B Virus**
**\*\*\*HCV: Hepatitis C Virus**
**\*\*\*\*COPD: Chronic Obstructive Pulmonary Disease**
**\*\*\*\*\*SD: Standard Deviation**

HIV coinfection was present for 16 persons (5%), HBV coinfection for 12 (4%), and HCV coinfection for 6 (2%). Alcohol use disorder and drug use disorder was diagnosed for 8 (3%) and 13 (4%) persons, respectively. The mean modified Charlson score (i.e. excluding HIV) was 1.8. Incident drug-induced liver injury during the 12-month follow-up period occurred for 13 persons (4%). A total of 16 persons (5%) had the earliest TB-related claims more than 365 days before their index date, suggesting prior or recurrent TB episodes.

### Total All Cause Healthcare Costs

Median total 12-month all-cause healthcare costs from the index date were $8,075 (IQR $3,001–$25,276) in 2018 USD (Table 2). Outpatient services accounted for the largest share of median cost ($4,717), followed by pharmacy ($1,254) and inpatient services ($0 overall; $36,785 among the 44 patients [14.5%] with at least one admission). Among admitted patients, median length of stay was 6.5 days (IQR 3–19 days). An additional 58 patients (19.1%) had at least one ED visit during the 12-month window. Among all 303 patients, median TB-specific medication costs (defined as pharmacy costs attributable to the four first-line antitubercular drugs (isoniazid, rifampin, pyrazinamide, and ethambutol)) were $310 per patient over the 12-month follow-up period, representing approximately 10% of total pharmacy costs and 2% of total healthcare costs.

**Table 2.**
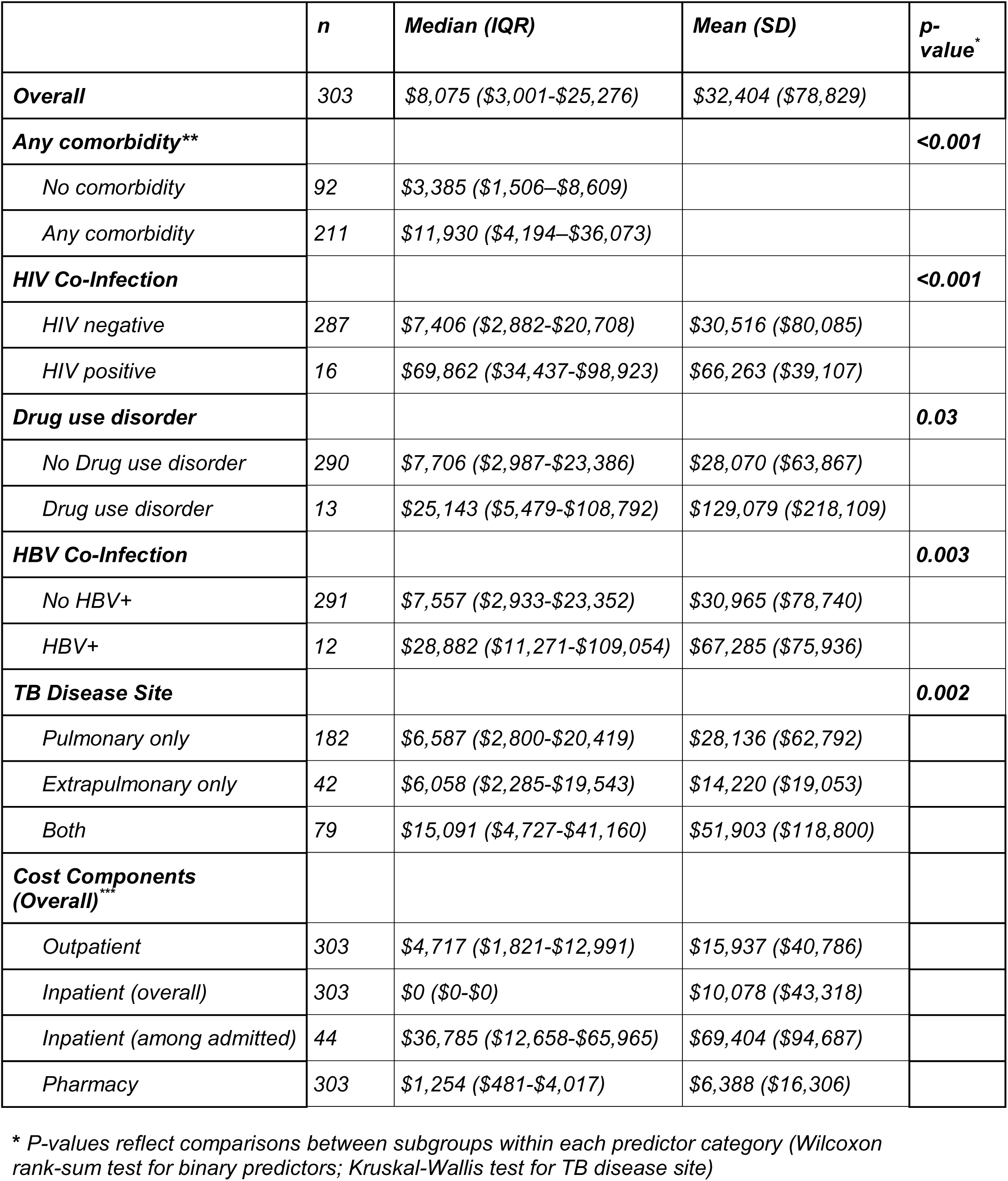
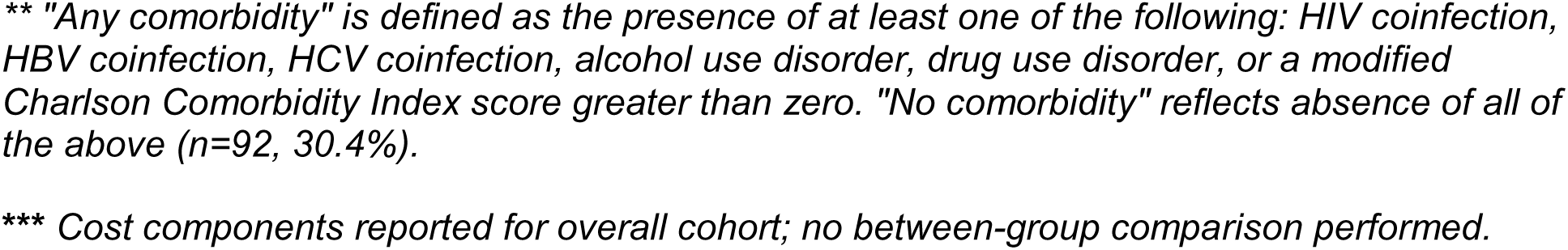
Unadjusted 12-Month All-Cause Costs Among Primary Cohort.

|  | *n* | *Median (IQR)* | *Mean (SD)* | *p-value\** |
| --- | --- | --- | --- | --- |
| **Overall** | 303 | \$8,075 (\$3,001-\$25,276) | \$32,404 (\$78,829) | |
| **Any comorbidity\*\*** |  |  |  | **<0.001** |
| No comorbidity | 92 | \$3,385 (\$1,506–\$8,609) | | |
| Any comorbidity | 211 | \$11,930 (\$4,194–\$36,073) | | |
| **HIV Co-Infection** |  |  |  | **<0.001** |
| HIV negative | 287 | \$7,406 (\$2,882-\$20,708) | \$30,516 (\$80,085) | |
| HIV positive | 16 | \$69,862 (\$34,437-\$98,923) | \$66,263 (\$39,107) | |
| **Drug use disorder** |  |  |  | **0.03** |
| No Drug use disorder | 290 | \$7,706 (\$2,987-\$23,386) | \$28,070 (\$63,867) | |
| Drug use disorder | 13 | \$25,143 (\$5,479-\$108,792) | \$129,079 (\$218,109) | |
| **HBV Co-Infection** |  |  |  | **0.003** |
| No HBV+ | 291 | \$7,557 (\$2,933-\$23,352) | \$30,965 (\$78,740) | |
| HBV+ | 12 | \$28,882 (\$11,271-\$109,054) | \$67,285 (\$75,936) | |
| **TB Disease Site** |  |  |  | **0.002** |
| Pulmonary only | 182 | \$6,587 (\$2,800-\$20,419) | \$28,136 (\$62,792) | |
| Extrapulmonary only | 42 | \$6,058 (\$2,285-\$19,543) | \$14,220 (\$19,053) | |
| Both | 79 | \$15,091 (\$4,727-\$41,160) | \$51,903 (\$118,800) | |
| **Cost Components (Overall)\*\*\*** |  |  |  |  |
| Outpatient | 303 | \$4,717 (\$1,821-\$12,991) | \$15,937 (\$40,786) | |
| Inpatient (overall) | 303 | \$0 (\$0-\$0) | \$10,078 (\$43,318) | |
| Inpatient (among admitted) | 44 | \$36,785 (\$12,658-\$65,965) | \$69,404 (\$94,687) | |
| Pharmacy | 303 | \$1,254 (\$481-\$4,017) | \$6,388 (\$16,306) | |

\* P-values reflect comparisons between subgroups within each predictor category (Wilcoxon rank-sum test for binary predictors; Kruskal-Wallis test for TB disease site)
*\*\* "Any comorbidity" is defined as the presence of at least one of the following: HIV coinfection, HBV coinfection, HCV coinfection, alcohol use disorder, drug use disorder, or a modified Charlson Comorbidity Index score greater than zero. "No comorbidity" reflects absence of all of the above (n=92, 30.4%).*
*\*\*\* Cost components reported for overall cohort; no between-group comparison performed.*

### Factors Associated with Increased Costs

Unadjusted 12-month costs differed substantially by HIV status; median costs were $69,862 among HIV-coinfected persons, as opposed to $7,406 among HIV-negative persons (Wilcoxon p<0.001). Median costs were also higher among persons with drug use disorder ($25,143 vs $7,706, p=0.03), HBV co-infection ($28,882 vs $7,557, p=0.003), and combined pulmonary and extrapulmonary TB vs. pulmonary-only TB ($15,091 vs $6,587, p<0.001). In adjusted analysis, four factors were significantly associated with higher 12-month total healthcare costs (**Table 3**). HIV co-infection had the largest effect, with an adjusted cost ratio (aCR) of 4.7 (95% CI 2.09w– 10.56, p<0.001). Drug use disorder was associated with 3.2 times higher adjusted costs (95% CI 1.29–8.02, p=0.01). Combined pulmonary and extrapulmonary disease vs pulmonary-only disease was associated with a 1.6-fold increase in adjusted costs. The modified Charlson comorbidity score was associated with a 1.4-fold increase in total costs per point, reflecting consistent increases of underlying composite comorbidity burden on cost. HBV coinfection, incident DILI, and alcohol use disorder showed point estimates suggestive of higher costs, but did not reach statistical significance.

**Table 3.** Adjusted All-Cause Predictors of 12-Month Total Healthcare Costs Among Primary Cohort.

| Predictor* | aCR** (95% CI***) | p-value |
| --- | --- | --- |
| Human Immunodeficiency Virus coinfection | 4.70 (2.09-10.56) | <0.001 |
| Drug use disorder | 3.22 (1.29-8.02) | 0.013 |
| Pulmonary + extrapulmonary TB**** | 1.60 (1.02-2.49) | 0.040 |
| Extrapulmonary-only TB | 1.05 (0.60-1.81) | 0.872 |
| Charlson comorbidity score (per point) | 1.36 (1.25-1.48) | <0.001 |
| Incident Drug-Induced Liver Injury | 1.71 (0.67-4.39) | 0.265 |
| Hepatitis B Virus coinfection | 1.48 (0.57-3.82) | 0.419 |
| Alcohol use disorder | 0.80 (0.25-2.53) | 0.702 |

\* All predictors listed were included simultaneously in a single multivariable Gamma generalized linear model with log link. The model additionally adjusted for age category (18–34, 35–49, 50–64, 65+), sex, US census region, and index calendar year (not shown).
\*\* aCR: Adjusted cost ratio - Estimated using separate Gamma generalized linear models with log link for each cost component: total 12-month all-cause costs, outpatient costs, inpatient costs (among the 44 admitted patients only), and outpatient pharmacy costs.
\*\*\* A 95% confidence interval that excludes 1.00 indicates statistical significance at $p < 0.05$ .
\*\*\*\* TB disease site is modeled as a 3-level categorical variable; pulmonary-only TB is the reference category. Adjusted cost ratios reflect costs relative to patients with pulmonary-only disease.

### Components of Increased Costs

To identify where each predictor’s cost effect was concentrated within the continuum of care, we fit parallel models for outpatient costs, inpatient costs (among admitted patients), outpatient pharmacy costs, and odds of hospitalization (**Table 4**)

**Table 4.** Cost Components of All-Cause Cost Predictors Among Primary Cohort*.

| Predictor* | Total cost<br>aCR (95%<br>CI)** | Outpatient<br>aCR (95% CI) | Inpatient<br>(admitted)<br>aCR (95%<br>CI) | Pharmacy<br>aCR (95% CI) | Hospitalization<br>(OR)<br>aOR (95% CI) |
| --- | --- | --- | --- | --- | --- |
| Human Immunodeficiency Virus coinfection | **4.70 (2.09-10.56)** | 1.31 (0.67-2.58) | 1.39 (0.48-4.04) | **16.37 (7.24-37.03)** | **6.38 (1.92-21.17)** |
| Drug use disorder | **3.22 (1.29-8.02)** | 1.85 (0.86-3.97) | **3.48 (1.25-9.73)** | **3.67 (1.46-9.23)** | **4.40 (1.17-16.57)** |
| Pulmonary + extrapulmonary TB | **1.60 (1.02-2.49)** | **1.62 (1.11-2.35)** | 1.48 (0.74-2.97) | **1.58 (1.01-2.47)** | **2.29 (1.04-5.03)** |
| Extrapulmonary TB- only | 1.05 (0.60-1.81) | 1.18 (0.75-1.87) | 0.72 (0.16-3.22) | 0.91 (0.52-1.59) | 0.44 (0.09-2.10) |
| Charlson (per point) | **1.36 (1.25-1.48)** | **1.36 (1.27-1.46)** | 1.11 (0.97-1.27) | **1.34 (1.23-1.46)** | **1.22 (1.07-1.41)** |
| Incident Drug Induced Liver Injury | 1.71 (0.67-4.39) | 0.73 (0.33-1.59) | 1.41 (0.36-5.55) | 0.72 (0.28-1.86) | 1.70 (0.37-7.88) |
| Hepatitis B Virus coinfection | 1.48 (0.57-3.82) | 1.18 (0.53-2.60) | 1.17 (0.33-4.14) | 1.48 (0.57-3.84) | 1.19 (0.25-5.66) |
| Alcohol use disorder | 0.80 (0.25-2.53) | 0.96 (0.37-2.52) | 0.43 (0.09-2.10) | 0.73 (0.23-2.34) | 1.11 (0.17-7.24) |

*Bolded values represent those with p value <.05*
*\*\*aCR: Adjusted cost ratio - Estimated using separate Gamma generalized linear models with log link for each cost component: total 12-month all-cause costs, outpatient costs, inpatient costs (among the 44 admitted patients only), and outpatient pharmacy costs.*
*\*\*\*aOR: Adjusted odds ratio - Estimated using logistic regression.*
*All models adjusted for the same covariates as the primary model: HIV coinfection, drug use disorder, alcohol use disorder, incident DILI, modified Charlson Comorbidity Index (continuous), TB disease site (pulmonary only, extrapulmonary only, both), HBV coinfection, age category, sex, U.S. census region of patient residence, and calendar year of index date. Bolded values indicate p<0.05. Reference category for TB disease site is pulmonary-only disease.*

HIV co-infection’s effect on total cost was driven mostly by pharmacy costs, with an adjusted pharmacy cost ratio of 16.4 (95% CI 7.2–37.0, p<0.001). HIV was also associated with six times higher odds of hospitalization (OR 6.38, 95% CI 1.92–21.17, p=0.002), but its effects on both outpatient costs and inpatient costs among hospitalized persons were smaller and not statistically significant.

Amongst persons with TB disease and drug use disorder, a different pattern emerged. Drug use disorder had significant effects on inpatient costs among hospitalized persons (aCR 3.48, p=0.024), pharmacy costs (aCR 3.67, p=0.006), and increased odds of hospitalization (OR 4.40, p=0.028), but not on outpatient costs.

Patients with combined extrapulmonary and pulmonary TB were significantly more likely to have higher outpatient (aCR 1.62, p=0.012) and pharmacy costs (aCR 1.58, p=0.048), as well as higher odds of hospitalization (OR 2.29, p=0.040) than persons with only pulmonary TB.

Modified Charlson score showed proportional effects on outpatient costs (aCR 1.36 per point), pharmacy costs (aCR 1.34 per point), inpatient costs among hospitalized persons (aCR 1.18 per point), and odds of hospitalization (OR 1.23 per point).

### Sensitivity Analyses

Primary findings were consistent across multiple sensitivity analyses (**Appendix B**). When we used a case definition requiring only one TB ICD code, all major effect estimates changed by less than 5%. Excluding the four persons above the 99th percentile for total cost attenuated effects modestly, but preserved direction and statistical significance for HIV, drug use disorder, Charlson score, and disseminated TB.

Using a 15-month expanded cost window (90 days pre-index through 365 days post-index) attenuated all effect estimates as expected when capturing additional non-TB attributable baseline healthcare use.

## DISCUSSION

Prior studies estimating TB care costs among the commercially insured have focused primarily on inpatient expenditure. Owusu et al.^4^ found average hospitalization expenditure per hospitalized person was $33,085, which is roughly in line with our estimate of $36,785 among hospitalized persons. The current analysis extends this work by characterizing total all-cause healthcare costs across outpatient, inpatient, and pharmacy settings, which has been infrequently described. Mean total 12-month all-cause costs of $32,404 per patient represent a higher economic burden than previously recognized from hospitalization-only analyses, reflecting the substantial outpatient and pharmacy costs that accumulate over the full course of TB treatment and comorbidity management. Applied to the approximately 23% of U.S. TB cases estimated to receive commercially insured care,^19^ our per-person cost estimate implies a national annual burden of roughly $71.2 million in all-cause healthcare expenditure among commercially insured TB persons diagnosed with TB. This figure is likely an underestimate, given that our 12-month window does not capture diagnostic-phase costs preceding treatment confirmation. As TB case counts in the U.S. have increased for four consecutive years and now exceed more than ten thousand cases annually,^20^ the commercial sector cost burden is likely growing, underscoring the importance of identifying and addressing the key drivers documented here.

This decision to use all-cause healthcare costs rather than TB-attributable costs was made to ensure that costs of coinfection management, comorbidity care, and treatment-related complications were included in our analysis on costs for the private payer system. Restricting to TB-only costs would have underestimated the true burden of TB treatment costs. This approach also carries a risk of linking costs for unrelated chronic conditions to TB. We mitigated this by including a modified Charlson Comorbidity Index (CCI) in our multivariable models. By adjusting for baseline comorbidity burden, our analysis delineates the incremental cost burden specifically associated with TB and its most significant clinical drivers, as opposed to the costs of persons’ pre-existing, non-TB-related healthcare needs. Our 12-month post-index cost window is a conservative measure of the full TB cost episode. In the 90 days prior to the index date, 90% of persons had at least one TB ICD code, reflecting diagnostic workup that precedes confirmation of diagnosis. The 15-month sensitivity analysis demonstrates that incorporating this pre-index period attenuates effect sizes but preserves directional findings.

Our most consistent finding across sensitivity analyses was that HIV-TB co-infection significantly increases costs for persons with TB disease, primarily through pharmacy expenditures. ART accounts for the majority of HIV-related expenditure, and list prices for first-line regimens are high and rising.^21^ Net public-sector ART costs are substantially lower due to Medicaid rebates and 340B pricing,^22^ so the pharmacy-driven HIV differential we observed is consistent with, though not direct evidence of, higher net ART expenditure among the commercially insured.

In addition, we observed the effect of drug use disorder on costs operated primarily through increased inpatient and pharmacy expenditures, rather than outpatient expenditures. This is consistent with existing evidence that drug use disorder increases complications through increasing odds of hospitalization, treatment failure, and treatment duration.^7^

We also observed that persons with claims for both pulmonary and extra-pulmonary TB had significantly higher costs than those with claims for pulmonary-only TB. Although this achieved marginal significance, it is in line with the evidence that hospitalization expenditures for some forms of extra-pulmonary TB were significantly higher than for pulmonary TB^4^ and extends this finding to both outpatient and pharmacy costs.

The modified Charlson Comorbidity Score also showed a strong, graded effect, reinforcing a recurring theme in chronic disease cost analyses that underlying comorbidity burden compounds expense across all care settings.^23^ Each additional Charlson point added roughly 36% to total costs, with effects distributed across all 3 cost components.

Several limitations in this study should be highlighted. First, the Marketscan database represents a convenience sample of employer-sponsored health plans and is not nationally representative. The MarketScan database cohort skews towards employed, working adults^9^, which is a group with relatively low TB prevalence.^20^ Second, our TB case definition systematically excluded persons whose TB care was largely managed outside the commercial sector. The primary objective of this study, however, was to identify predictors of cost variation rather than to estimate population-level costs, and analyses using the more sensitive definition (requiring only one TB ICD code) yielded consistent findings.

Comorbidity ascertainment relied on ICD codes in the six-month pre-index lookback window using a previously published and validated approach. Persons whose comorbidities were managed outside the commercial sector or not coded during the lookback period will be assigned lower Charlson scores than their true experience. This potential misclassification biases the Charlson comorbidity coefficient toward the null, meaning our estimate of 1.36-fold higher costs per Charlson point is likely a conservative lower bound of the true effect of comorbidity on costs. Notably, this same limitation applies to alcohol use disorder and HBV coinfection, whose non-significant findings may partly reflect measurement error rather than a true absence of effect.

Because the database begins in January 2013, patients with early index dates could not have prior claims fully verified, likely leaving a small number of recurrent cases in the primary cohort. The consistency of the recurrence-excluded sensitivity analysis suggests this does not materially affect results.

## IMPLICATIONS FOR POLICY AND PRACTICE

- As TB diagnoses rise and commercial insurance covers an increasing share of persons in the U.S., these findings have direct implications for payers and health care systems. HIV and HBV coinfection are identifiable before TB diagnosis and commercial insurers could reduce downstream costs by incentivizing proactive screening for these conditions among high-risk members. This can enable earlier TB diagnosis when treatment is less complex and less costly.
- Our finding that costs rose with comorbidity burden reinforces existing recommendations for targeted latent TB infection (LTBI) testing and treatment in these groups, where progression risk and downstream cost are both concentrated.
- Emerging U.S. evidence suggests LTBI prevalence is elevated in populations with specific chronic diseases, such as HBV and HCV. Among adults with chronic HBV infection who underwent LTBI testing, coinfection prevalence was more than two-fold higher than adults without chronic HBV.^24^ Persons with diabetes have also been found to have higher LTBI prevalence.^25^
- Because our cohort comprised persons with TB disease, we cannot directly assess LTBI prevalence and costs. However, the cost gradient observed across Charlson Comorbidity Index scores suggests that when TB disease does develop in medically complex persons predisposed to LTBI, resulting expenditures are disproportionately high.

## CONCLUSION

In commercially insured U.S. adults with TB disease, total 12-month healthcare costs averaged $32,404 per patient and were independently driven by HIV coinfection, drug use disorder, combined pulmonary/extrapulmonary TB, and comorbidity burden. Preventing progression to TB in these populations may avert significant disease and economic costs. Future research should validate these findings in larger datasets including Medicare claims and linked surveillance data that permit race/ethnicity-stratified and TB-attributable cost analyses. These findings also highlight the need for LTBI testing and treatment in persons with comorbidities. As U.S. TB incidence increases for the fourth consecutive year, understanding and addressing the commercial sector cost burden is an urgent public health and policy priority.

## Conflicts of Interest and Source of Funding

The authors have no conflicts of interest to declare. No funding was used to support this study.

## Data Availability Statement

The data that support the findings of this study were obtained under license from the Merative™ MarketScan® Commercial Claims Database and are not publicly available. Access was provided through the UCSF Institutional Data Access program. Restrictions apply to the availability of these data, which were used under license for the current study.

## Acknowledgments

The authors thank the University of California, San Francisco Institute for Health Policy Studies and School of Medicine for providing access to the Merative MarketScan Commercial Claims and Encounters Database through the UCSF Institutional Data Access program.

## Implications for Policy and Practice

Comorbidity burden (particularly HIV coinfection and drug use disorder) is a major driver of healthcare costs among commercially-insured persons with tuberculosis, supporting targeted screening and early intervention in these high-risk groups.

## Notes

### Competing Interest Statement

The authors have declared no competing interest.

### Summary of Updates

Author order updated to reflect the manuscript

