## Appendix for "Predictors of Healthcare Costs among Commercially-Insured Persons with Tuberculosis in the United States, 2013 to 2018"

### Appendix A: Charlson Comorbidity Index* — ICD-9-CM and ICD-10 Coding Algorithms

*Source: Quan H, Sundararajan V, Halfon P, et al. Coding Algorithms for Defining Comorbidities in ICD-9-CM and ICD-10 Administrative Data. Medical Care. 2005;43(11):1130-1139.*

| | Comorbidity | Weight | ICD-9-CM Codes (Quan 2005 Enhanced) | ICD-10 Codes (Quan 2005) |
|---|---|---|---|---|
| 1 | Myocardial infarction | 1 | 410.x, 412.x | I21.x, I22.x, I25.2 |
| 2 | Congestive heart failure | 1 | 398.91, 402.01, 402.11, 402.91, 404.01, 404.03, 404.11, 404.13, 404.91, 404.93, 425.4–425.9, 428.x | I09.9, I11.0, I13.0, I13.2, I25.5, I42.0, I42.5–I42.9, I43.x, I50.x, P29.0 |
| 3 | Peripheral vascular disease | 1 | 093.0, 437.3, 440.x, 441.x, 443.1–443.9, 447.1, 557.1, 557.9, V43.4 | I70.x, I71.x, I73.1, I73.8, I73.9, I77.1, I79.0, I79.2, K55.1, K55.8, K55.9, Z95.8, Z95.9 |
| 4 | Cerebrovascular disease | 1 | 362.34, 430.x–438.x | G45.x, G46.x, H34.0, I60.x–I69.x |
| 5 | Dementia | 1 | 290.x, 294.1, 331.2 | F00.x–F03.x, F05.1, G30.x, G31.1 |
| 6 | Chronic pulmonary disease | 1 | 416.8, 416.9, 490.x–505.x, 506.4, 508.1, 508.8 | I27.8, I27.9, J40.x–J47.x, J60.x–J67.x, J68.4, J70.1, J70.3 |
| 7 | Rheumatic disease | 1 | 446.5, 710.0–710.4, 714.0–714.2, 714.8, 725.x | M05.x, M06.x, M31.5, M32.x–M34.x, M35.1, M35.3, M36.0 |
| 8 | Peptic ulcer disease | 1 | 531.x–534.x | K25.x–K28.x |
| 9 | Mild liver disease | 1 | 070.22, 070.23, 070.32, 070.33, 070.44, 070.54, 070.6, 070.9, 570.x, 571.x, 573.3, 573.4, 573.8, 573.9, V42.7 | B18.x, K70.0–K70.3, K70.9, K71.3–K71.5, K71.7, K73.x, K74.x, K76.0, K76.2–K76.4, K76.8, K76.9, Z94.4 |

| | | | | |
|---|---|---|---|---|
| 10 | Diabetes without chronic complication | 1 | 250.0–250.3, 250.8, 250.9 | E10.0, E10.1, E10.6, E10.8, E10.9, E11.0, E11.1, E11.6, E11.8, E11.9, E12.0, E12.1, E12.6, E12.8, E12.9, E13.0, E13.1, E13.6, E13.8, E13.9, E14.0, E14.1, E14.6, E14.8, E14.9 |
| 11 | Diabetes with chronic complication | 2 | 250.4–250.7 | E10.2–E10.5, E10.7, E11.2–E11.5, E11.7, E12.2–E12.5, E12.7, E13.2–E13.5, E13.7, E14.2–E14.5, E14.7 |
| 12 | Hemiplegia or paraplegia | 2 | 334.1, 342.x, 343.x, 344.0–344.6, 344.9 | G04.1, G11.4, G80.1, G80.2, G81.x, G82.x, G83.0–G83.4, G83.9 |
| 13 | Renal disease | 2 | 403.01, 403.11, 403.91, 404.02, 404.03, 404.12, 404.13, 404.92, 404.93, 582.x, 583.0–583.7, 585.x, 586.x, 588.0, V42.0, V45.1, V56.x | I12.0, I13.1, N03.2–N03.7, N05.2–N05.7, N18.x, N19.x, N25.0, Z49.0–Z49.2, Z94.0, Z99.2 |
| 14 | Any malignancy (incl. lymphoma and leukemia, except non-melanoma skin) | 2 | 140.x–172.x, 174.x–195.8, 200.x–208.x, 238.6 | C00.x–C26.x, C30.x–C34.x, C37.x–C41.x, C43.x, C45.x–C58.x, C60.x–C76.x, C81.x–C85.x, C88.x, C90.x–C97.x |
| 15 | Moderate or severe liver disease | 3 | 456.0–456.2, 572.2–572.8 | I85.0, I85.9, I86.4, I98.2, K70.4, K71.1, K72.1, K72.9, K76.5, K76.6, K76.7 |
| 16 | Metastatic solid tumor | 6 | 196.x–199.x | C77.x–C80.x |

| | | | | |
|---|---|---|---|---|
| 17 | AIDS/HIV | 6 | Excluded from modified Charlson | Excluded from modified Charlson |

**Charlson Score Comorbidity Calculation - If any ICD-9 or ICD-10 code in the patient's 6-month pre-index lookback window includes the following diagnosis, add the corresponding number of points for each. This definition is nearly identical to the one specified by Quan et al 2005, but removes the HIV/AIDS component because we are using HIV/AIDS as a separate predictor variable in our GLM Log Link model.*

### Appendix B: Sensitivity Analyses of Adjusted Cost Ratios for Predictors of 12-Month All-Cause Healthcare Costs, Evaluating Robustness to Cost Window Definition, Outlier Exclusion, and Exclusion of Potential Recurrent TB Diagnoses

| **Sensitivity Analysis** | **Human Immunodefeciency Virus**<br><br>aCR* (95% CI) | **Drug use**<br><br>aCR (95% CI) | **Charlson**<br><br>aCR (95% CI) | **Extrapulmonary and pulmonary TB**<br><br>aCR (95% CI) | **Hepatitis B Virus**<br><br>aCR (95% CI) |
|---|---|---|---|---|---|
| **Primary model (12-month window, N=303)**** | ***4.70 (2.09-10.56)*** | ***3.22 (1.29-8.02)*** | ***1.36 (1.25-1.48)*** | ***1.60 (1.02-2.49)*** | 1.48 (0.57-3.82) |
| **15-month expanded window (N=303)***** | ***3.00 (1.61-5.61)*** | 2.00 (0.98-4.05) | ***1.26 (1.18-1.35)*** | 1.35 (0.95-1.90) | 1.18 (0.57-2.46) |
| **Outliers excluded (top 1%, N=299)****** | ***4.76 (2.21-10.24)*** | ***3.01 (1.17-7.73)*** | ***1.32 (1.21-1.43)*** | ***1.60 (1.05-2.46)*** | 1.60 (0.65-3.94) |
| **Recurrence cases excluded (gap >365 days, N=287)******* | ***4.47 (1.95-10.20)*** | ***2.96 (1.15-7.63)*** | ***1.37 (1.26-1.50)*** | ***1.59 (1.01-2.48)*** | 1.20 (0.45-3.22) |

**aCR: Adjusted cost ratio - Estimated using separate Gamma generalized linear models with log link for 12-Month All-Cause Healthcare Costs*

***Primary model: 12-month post-index cost window, full cohort (N=303).*

****15-month expanded window: costs captured from 90 days pre-index through 365 days post-index.*

*****Outliers excluded: four persons with total costs exceeding the 99th percentile ($412,922) removed.*

******Recurrence excluded: 16 persons with earliest TB-related claims more than 365 days before the index date removed.*
